# Complementary Value of Transthoracic Echocardiography and Transcranial Doppler for Screening in Cryptogenic Stroke

**DOI:** 10.64898/2026.01.08.26343740

**Authors:** Ken Oshima, Yasuhide Mochizuki, Keita Mizuma, Tetsuhito Nohara, Ayako Miki, Mamiko Yamada, Ayaka Oda, Yumi Yamamoto, Sakiko Gohbara, Saaya Ichikawa-Ogura, Rumi Hachiya, Eiji Toyosaki, Hiroto Fukuoka, Hidetomo Murakami, Naoki Uchida, Toshiro Shinke

## Abstract

**Background:** The optimal noninvasive screening strategy for detecting patent foramen ovale (PFO) in patients with cryptogenic stroke (CS) remains uncertain. Although transthoracic echocardiography (TTE) and transcranial Doppler (TCD) are widely used, whether combining both modalities improves diagnostic performance has not been fully established.

**Methods:** Among 432 consecutive CS patients, 399 underwent collaborative screening with both TTE and TCD bubble tests performed by a multidisciplinary Heart–Brain Team, followed by transesophageal echocardiography (TEE) as the reference standard. Bubble tests were conducted at rest and during the Valsalva maneuver (VM). Diagnostic performance, concordance between modalities, and incremental value were evaluated using receiver operating characteristic analysis, Cohen’s kappa statistics, and sequential logistic regression models.

**Results:** TEE confirmed PFO in 156 patients (39.1%). Both TTE and TCD demonstrated significantly higher diagnostic accuracy during VM than at rest, with no significant difference in area under the curve between modalities under VM. Sequential logistic regression showed a significant incremental increase in predictive value when TCD during VM was added to TTE during VM (χ² increase from 271.4 to 297.2; p<0.0001). Although overall agreement between TTE and TCD during VM was substantial (κ=0.63), 54 patients (14%) showed discordant results, among whom 15 (28%) had TEE-confirmed PFO. Applying an “OR” rule (positive if either test was positive) significantly improved sensitivity compared with either modality alone, at the expense of modestly reduced specificity.

**Conclusions:** Dual screening with TTE and TCD during VM within a Heart–Brain Team framework significantly enhances sensitivity for PFO detection and reduces missed diagnoses in patients with CS. An “OR” rule interpretation represents a practical and clinically effective screening strategy.

## Introduction

Cryptogenic stroke (CS) refers to cerebral infarction of undetermined etiology that cannot be explained by established mechanisms even after extensive diagnostic work-up, accounting for approximately 25% of all ischemic strokes^1^. Autopsy studies have demonstrated that a patent foramen ovale (PFO) is present in about 20–25% of the general population^2–4^, whereas it is detected in 40–50% of patients with CS^5^. In younger individuals, high-risk morphological features such as atrial septal aneurysm are more frequently observed, suggesting a particularly strong association between PFO and CS in this age group^6^. Several randomized clinical trials have demonstrated that appropriate transcatheter closure of PFO is superior to medical antithrombotic therapy in preventing recurrent ischemic stroke^7–10^. Therefore, accurately identifying patients in whom PFO represents the likely cause of CS is a clinically crucial step that may directly influence prognosis. However, in patients with CS and PFO, a definite venous thromboembolic source is rarely identified^11, 12^, and paradoxical embolism often remains a presumptive rather than a proven diagnosis. Consequently, the accuracy and quality of noninvasive diagnostic tools used to detect PFO and confirm right-to-left shunt (RLS) are of paramount importance in contemporary clinical practice. Contrast studies using microbubbles—either by transthoracic echocardiography (TTE) or transcranial Doppler (TCD)—are considered valuable noninvasive screening modalities for detecting RLS through a PFO. In patients with positive findings, transesophageal echocardiography (TEE) is recommended for definitive confirmation of PFO, characterization of its morphology, and detailed assessment of surrounding structures. However, each modality has inherent advantages and limitations: TTE tends to have relatively low sensitivity, whereas TCD offers higher sensitivity but may yield false-positive results due to the detection of extracardiac shunts such as pulmonary arteriovenous malformations^13–17^.

In recent years, the concept of the Heart–Brain Team (HBT) has gained increasing recognition of stroke management, emphasizing the importance of close collaboration between cardiologists and neurologists for the evaluation and treatment of patients with CS, including paradoxical embolism through a PFO^18–20^. However, the choice of screening modality may depend on which specialty first evaluates the patient, potentially leading to discrepant diagnostic outcomes. It remains uncertain whether screening with either TTE or TCD alone is sufficient, or whether a multidisciplinary approach combining both modalities can further enhance diagnostic accuracy. Therefore, this study aimed to investigate whether dual screening with TTE and TCD—performed collaboratively by cardiologists and stroke specialists within an HBT—improves the diagnostic performance for detecting PFO during the initial evaluation of patients with CS.

## Methods

### Establishment of a multidisciplinary Heart–Brain Team screening and decision-making framework

At our institution, stroke specialists traditionally performed the initial screening for PFO using TCD and TEE. Since April 2019, however, bubble screening for suspected paradoxical embolism in patients with CS has been performed jointly by at least two board-certified cardiologists and two stroke neurologists using both TTE and TCD. TEE has also been performed collaboratively by both departments to comprehensively evaluate intracardiac thrombus, PFO, valvular diseases, and aortic atherosclerotic plaques. In addition, beginning in the same period, a multidisciplinary case conference involving cardiologists, neurologists, neurosurgeons, and pediatric cardiologists has been held monthly to discuss each case, determine therapeutic indications, and develop individualized treatment strategies.

### Study population

From October 2019 to August 2025, 432 consecutive patients who were diagnosed at the Stroke Center of Showa Medical University Hospital by stroke specialists as having a high likelihood of cardioembolic stroke or as potentially having CS were included. In addition, patients who were referred from other institutions during the same period for evaluation of suspected PFO-related CS were included. Among these, 33 patients in whom either TTE or TCD were not performed or could not be adequately evaluated for any reason were excluded from this study. Therefore, the final study population thus comprised 399 patients who underwent both TTE and TCD bubble studies performed collaboratively by cardiologists and neurologists within the Brain–Heart Team, and in whom the presence or absence of PFO was subsequently confirmed by TEE. This study was approved by the local ethics committee of Showa Medical University Research Administration Centre (approval number: 2025-0407) and conducted in accordance with the Declaration of Helsinki. And it was also registered with the University Hospital Medical Information Network (UMIN 000060079). Opt-out informed consent procedure was applied for the use of participant data for research purposes. This consent procedure was reviewed and approved by the Showa Medical University Research Administration Centre, approval number 2025-0407, date of decision December 1, 2025.

### Contrast Test in Ultrasound Examinations and Valsalva Maneuver Procedure

Before the examination, all patients were trained to perform the Valsalva maneuver (VM). For patients who had difficulty generating sufficient abdominal pressure, manual abdominal compression was applied to assist the maneuver. Patients were positioned supine, and a 20-gauge intravenous catheter was placed in the right median cubital vein. The catheter was connected to an extension tube and then linked to three Luer-Lock syringes via a three-way stopcock. A mixture of 8 mL of normal saline, 1 mL of air, and 1 mL of the patient’s own blood was agitated vigorously between two syringes at least 10 times to create a microbubble suspension (Figure 1)^18, 21, 22^. This mixture was then immediately injected intravenously, followed by a 10 mL saline flush as a bolus push. During the bubble test, the patient performed the VM by maintaining inspiration and increasing intra-abdominal pressure for approximately 10 seconds. Microbubbles were injected just before release of the strain phase of VM. Bubble test on TTE was performed twice (at rest and during VM) by cardiology specialists. RLS at rest was regarded as a marker of high-risk morphology. Contrast microbubbles in the left atrium/ventricle within three cardiac cycles were counted and graded: negative (0), small (1–20), medium (> 20), and large (complete opacification) (Table 1). In TCD evaluation using the same sector probe as for TTE, the right middle cerebral artery was insonated through the temporal acoustic window. If adequate visualization could not be achieved, the vertebral artery was examined via a transforaminal (foramen magnum) approach using color Doppler imaging. During both resting and VM conditions, the contrast agent was intravenously injected, and micro-embolic signals (MES) were monitored and counted over a 30-second period. The detection of one or more MES was regarded as positive. The grading of shunt severity was determined by stroke specialists: negative: 0 signals; small: 1–10 signals; medium: > 10 signals without a curtain pattern; large: curtain-like appearance (uncountable) (Table 1)^23^.

**Figure 1.**
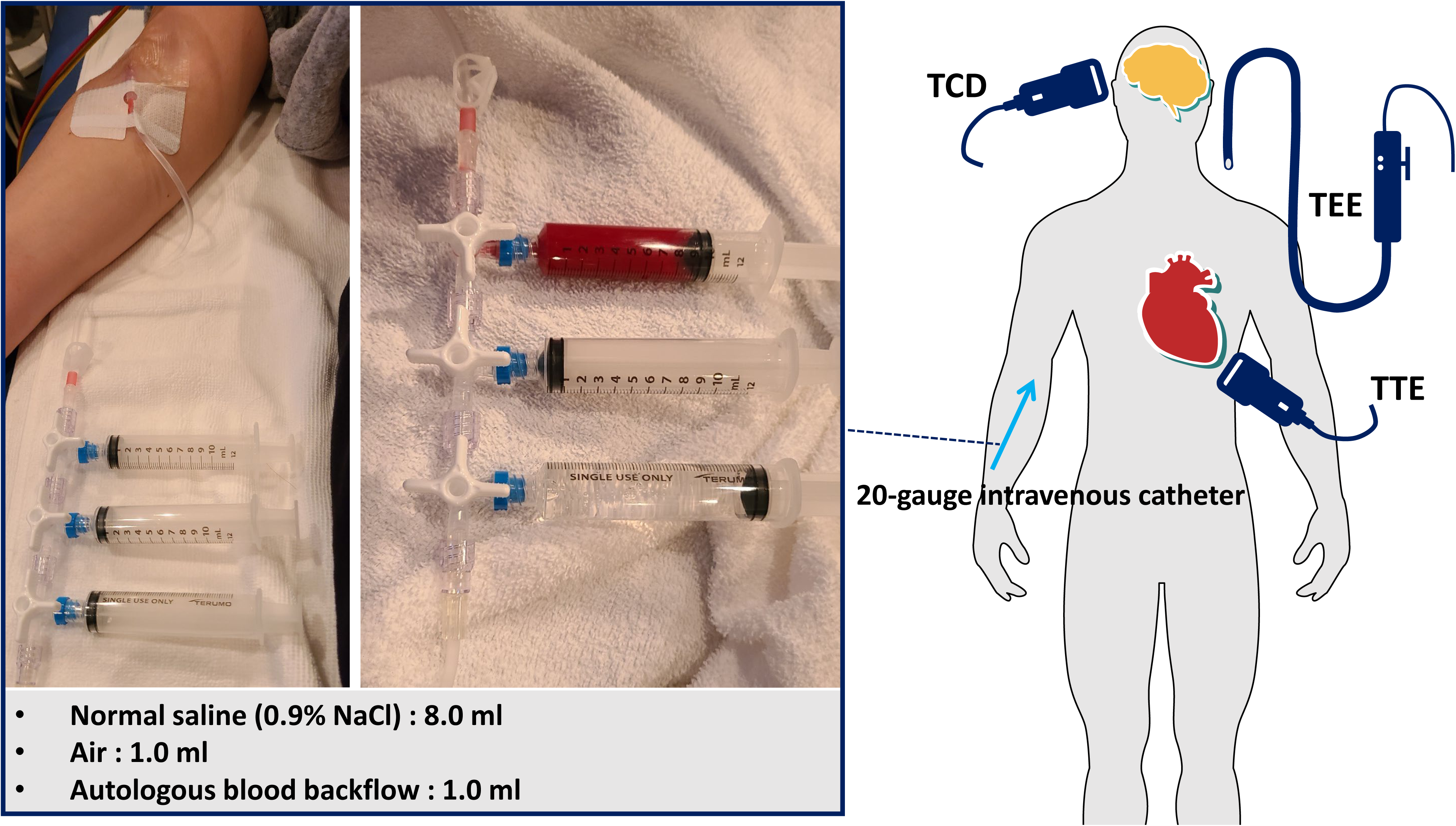
Preparation of agitated saline contrast for bubble testing. A 20-gauge intravenous catheter was placed in the right median cubital vein and connected via an extension tube to three Luer-Lock syringes through a three-way stopcock. A mixture of 8 mL normal saline, 1 mL air, and 1 mL autologous blood was agitated between two syringes at least 10 times to generate microbubbles for contrast studies during TTE, TCD, and TEE examinations. TTE = transthoracic echocardiography, TCD = transcranial Doppler, TEE = transesophageal echocardiography

**Table 1.** Grading of Right-to-Left Shunt by transthoracic echocardiography and transcranial Doppler.

| **TTE** | **TCD** | **Shunt Grade** |
| --- | --- | --- |
| Microbubbles counted in the left atrium and ventricle within 3 cardiac cycles after release of the VM | MES counted for 30 seconds in the right middle cerebral artery after release of the VM |  |
| Negative: 0 bubbles | Negative: 0 signals | **negative** |
| • 1–20 bubbles | • 1–10 MES | **small** |
| • $\geq 20$ bubbles | • $> 10$ MES without curtain pattern | **medium** |
| • Complete opacification of left heart | • Curtain-like (uncountable) | **large** |

TTE = transthoracic echocardiography; TCD = transcranial Doppler; VM = Valsalva Maneuver; MES = Micro- embolic signals

### Transesophageal echocardiography

Under the supervision of experienced cardiologists, a systematic assessment of valvular disease and cardiac hemodynamics was performed. The left atrial appendage and left atrium were examined for the presence and severity of spontaneous echo contrast and thrombus formation. Additionally, atherosclerotic plaques in the descending and aortic arch segments were evaluated. The interatrial septum was visualized, and during resting conditions, the contrast agent was injected to count the number of microbubbles appearing in the left atrium and left ventricle within three cardiac cycles. For the VM, manual abdominal compression was applied to augment intrathoracic pressure. The contrast agent was injected simultaneously with the release of the VM, and microbubbles appearing in the left atrium within three cardiac cycles after release were counted. When microbubbles were observed passing through a PFO, a RLS was diagnosed as positive.

### Statistical analysis

Statistical analyses were performed using data from patients with complete datasets. Continuous variables are presented as median (interquartile range), and categorical variables are expressed as numbers (percentages). Between-group differences in continuous variables were assessed using the Wilcoxon rank-sum test, and proportions were compared using Fisher’s exact test.

To evaluate the incremental value of adding TTE and TCD performed under VM to resting measurements for predicting the presence of PFO, a sequential logistic regression model was constructed. The presence of a PFO confirmed by TEE (i.e., RLS through the PFO) was defined as the dependent variable, and the results of each screening test (TTE and TCD) were entered as independent variables. First, a model including both TTE (rest) and TCD (rest) was used to calculate the chi-square statistic. Subsequently, additional variables were sequentially entered (TTE [Valsalva], then TCD [Valsalva]) to evaluate the improvement in model performance. At each step, the likelihood ratio test was used to assess whether the increase in chi-square value was statistically significant, thereby quantifying the incremental predictive value obtained by sequentially adding TTE and TCD parameters. Statistical significance was set at p < 0.05. All statistical analyses were performed using MedCalc Version 23.3.7 (MedCalc Software, Mariakerke, Belgium).

## Results

### Patient Background

Baseline clinical characteristics of the study population are summarized in Table 2. A total of 399 patients were analyzed. The median age was 68.0 (interquartile range [IQR], 55.0-77.0) years, and 233 patients (58.3%) were male. The median Risk of Paradoxical Embolism (RoPE) score^24^ was 4.0 (IQR, 3.0 – 5.0). Deep vein thrombosis (DVT) was identified in 19 patients (4.8%). TEE detected PFO in 156 patients (39.1%). Patients with PFO were significantly younger (median 61.5 vs 73.0 years, p < 0.05), had higher BMI (22.9 vs 21.6 kg/m², p < 0.05), and higher RoPE scores (p < 0.05) compared with those without PFO. The prevalence of hypertension was lower, whereas cortical infarction and history of DVT/PE were more frequent in the PFO group (p < 0.05).

**Table 2.** Baseline Characteristics of the Study Population According to the Presence or Absence of PFO.

|  | All (n = 399) | PFO (+) (n = 156) | PFO (–) (n = 243) | p value* |
| --- | --- | --- | --- | --- |
| Age, years | 68.0 (55.0-77.0) | 61.5 (47.3-72.0) | 73.0 (60.0-80.0) | p < 0.001 |
| Male sex, n (%) | 233 (58.3) | 136 (87.2) | 97 (39.9) | p < 0.001 |
| BMI, kg/m<sup>2</sup> | 22.2 (20.1-24.8) | 22.9 (21.0-25.4) | 21.6 (19.7-24.5) | p = 0.011 |
| RoPE score, median (IQR) | 4.0 (3.0-5.0) | 4.0 (3.0-7.0) | 3.0 (2.0-4.0) | p < 0.001 |
| Hypertension, n (%) | 234 (58.8) | 80 (51.3) | 154 (63.4) | p = 0.022 |
| Diabetes mellitus, n (%) | 68 (17.1) | 22 (14.1) | 46 (18.9) | p = 0.223 |
| Previous stroke/TIA, n (%) | 79 (19.8) | 25 (16.1) | 54 (22.2) | p = 0.157 |
| Smoking, n (%) | 141 (35.4) | 60 (38.5) | 81 (33.3) | p = 0.334 |
| Cortical infarction, n (%) | 93 (23.4) | 68 (43.6) | 25 (10.2) | p < 0.001 |
| History of DVT or PE, n (%) | 19 (4.9) | 12 (7.7) | 7 (2.9) | p = 0.032 |

PFO = patent foramen ovale; BMI = body mass index; RoPE = risk of paradoxical embolism; IQR = interquartile range;
TIA = transient ischemic attack; DVT = deep vein thrombosis; PE = pulmonary embolism
Data are presented as median (interquartile range) or number (%). P values indicate comparisons between patients with and
without PFO (Wilcoxon rank-sum test or Fisher exact test, as appropriate). \*p < 0.05 was considered statistically significant.

### Diagnostic performance of TTE and TCD for detection of PFO

Diagnostic sensitivity and specificity for each shunt grade threshold on both TTE and TCD—under resting conditions and during VM—were calculated and are presented in Figure 2. For both physiological states, sensitivity was highest when a threshold of small or greater was used to define a positive study, albeit at the cost of the lowest specificity. Conversely, when the threshold was set at large, sensitivity was lowest while specificity was correspondingly highest.

**Figure 2.**
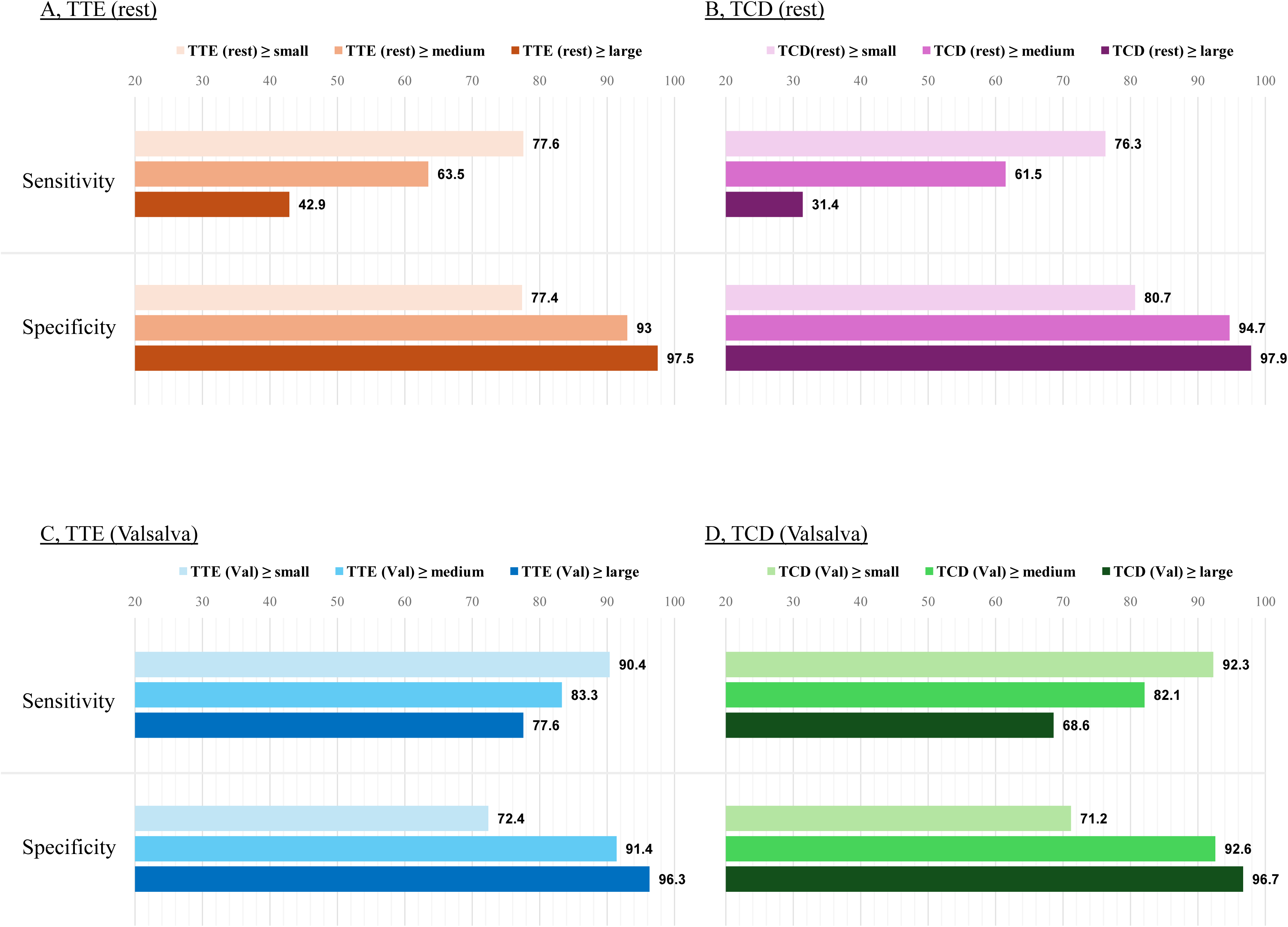
Comparison of sensitivity and specificity for PFO detection according to different shunt-grade cutoffs. Sensitivity and specificity for the detection of PFO according to different shunt-grade thresholds on transthoracic echocardiography (TTE) and transcranial Doppler (TCD). Analyses were performed under resting conditions (A, TTE; B, TCD) and during the Valsalva maneuver (C, TTE; D, TCD). Diagnostic performance was evaluated using three cutoff definitions: grade ≥ small, grade ≥ medium, and grade ≥ large. Across both modalities and physiological conditions, defining positivity as grade ≥ small yielded the highest sensitivity with the lowest specificity, whereas using grade ≥ large resulted in the lowest sensitivity and the highest specificity. TTE = transthoracic echocardiography, TCD = transcranial Doppler, TEE = transesophageal echocardiography

Among patients with TEE-confirmed PFO (shunt grade ≥ small), when positivity on both TTE and TCD was defined as shunt grade ≥ small, diagnostic accuracy parameters were as follows: TTE (rest), sensitivity 77.6%, specificity 77.4%; TCD (rest), sensitivity 76.3%, specificity 80.7%; TTE (Valsalva), sensitivity 90.4%, specificity 72.4%; and TCD (Valsalva), sensitivity 92.3%, specificity 71.2% (Table 3a). Comparison in ROC curve analysis revealed that both TTE and TCD with the VM exhibited significantly improved sensitivity and specificity, with AUCs significantly higher than those obtained at rest; however, no significant difference in AUC was observed between TTE and TCD under VM (Table 3b, Figure 3).

**Figure 3.**
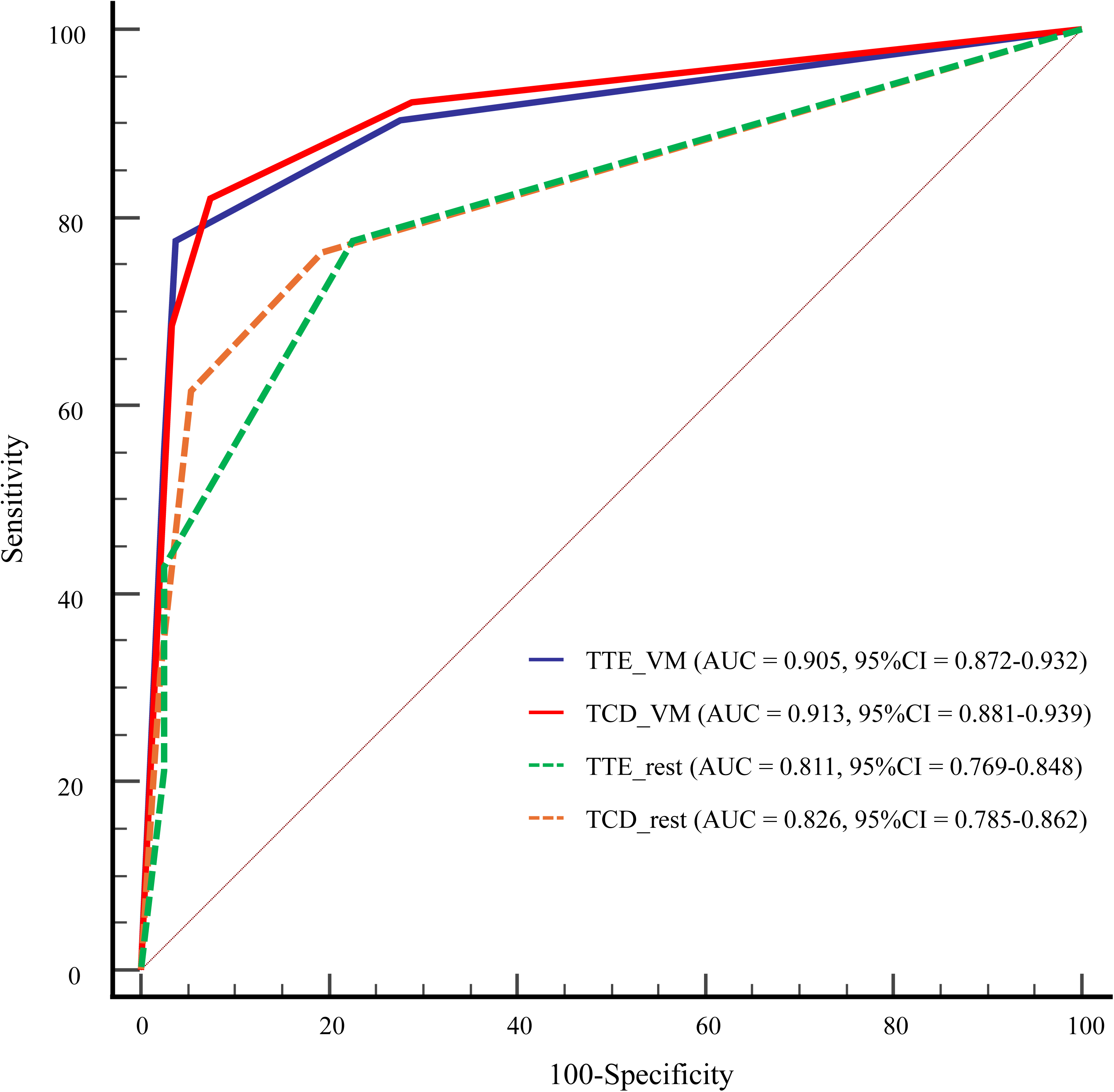
Receiver operating characteristic curves comparing diagnostic performance of TTE and TCD with and without Valsalva maneuver. Both TTE and TCD demonstrated significantly higher diagnostic accuracy under VM than at rest. The area under the curve (AUC)significantly increased from 0.811 (TTE-rest) and 0.826 (TCD-rest) to 0.905 (TTE-VM) and 0.913 (TCD-VM), respectively, with no significant difference between TTE and TCD during VM. TTE = transthoracic echocardiography, TCD = transcranial Doppler, VM = Valsalva maneuver

**Table 3a.**
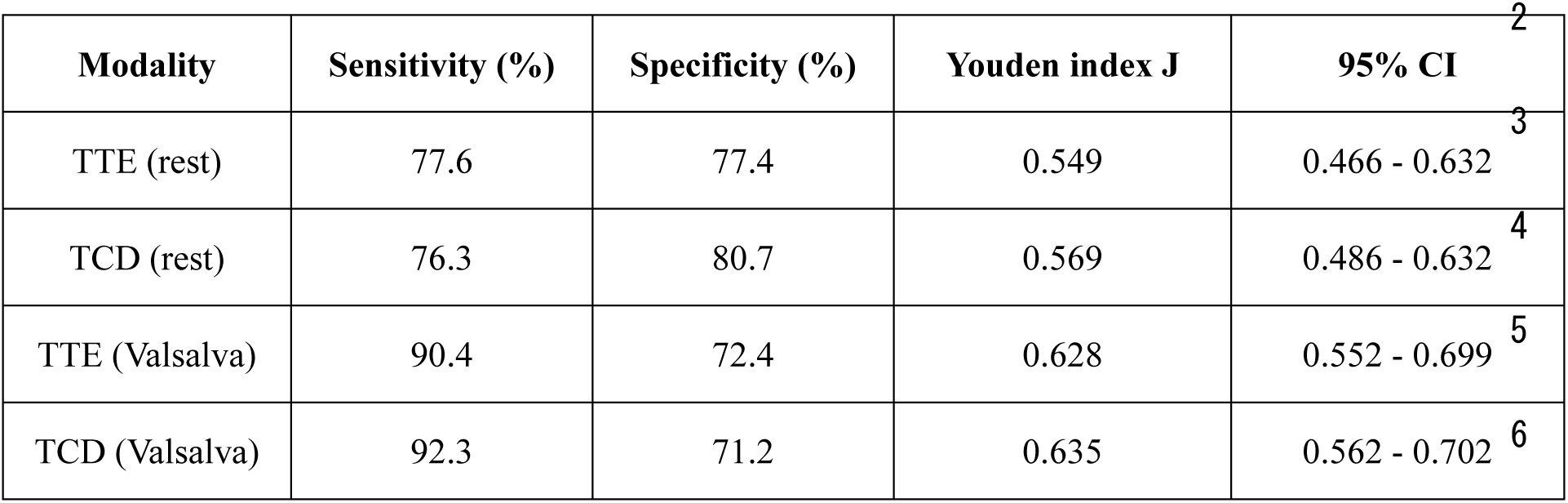
Diagnostic performance of TTE and TCD for detection of PFO.

**Table 3b.** Pairwise comparison of AUCs between TTE and TCD.

| Comparison | ΔAUC | 95% CI | z value | P value |
| --- | --- | --- | --- | --- |
| TTE (Valsalva) vs. TTE (Rest) | 0.094 | 0.057 – 0.131 | 4.94 | <0.0001 |
| TCD (Valsalva) vs. TCD (Rest) | 0.088 | 0.048 – 0.127 | 4.38 | <0.0001 |
| TTE (Valsalva) vs. TCD (Valsalva) | 0.009 | –0.023 – 0.040 | 0.52 | 0.600 |
| TTE (Valsalva) vs. TCD (Rest) | 0.079 | 0.037 – 0.121 | 3.69 | 0.0002 |
| TCD (Valsalva) vs. TTE (Rest) | 0.102 | 0.059 – 0.146 | 4.59 | <0.0001 |
| TCD (Rest) vs. TTE (Rest) | 0.015 | –0.023 – 0.053 | 0.77 | 0.445 |

PFO = patent foramen ovale; TTE = transthoracic echocardiography; TCD = transcranial Doppler; CI = confidence interval;
AUC = areas under the ROC curve

### Incremental predictive value of adding TCD to TTE

To assess the incremental predictive value of adding TCD under VM for identifying PFO, a sequential logistic regression model was constructed (Figure 4).

**Figure 4.**
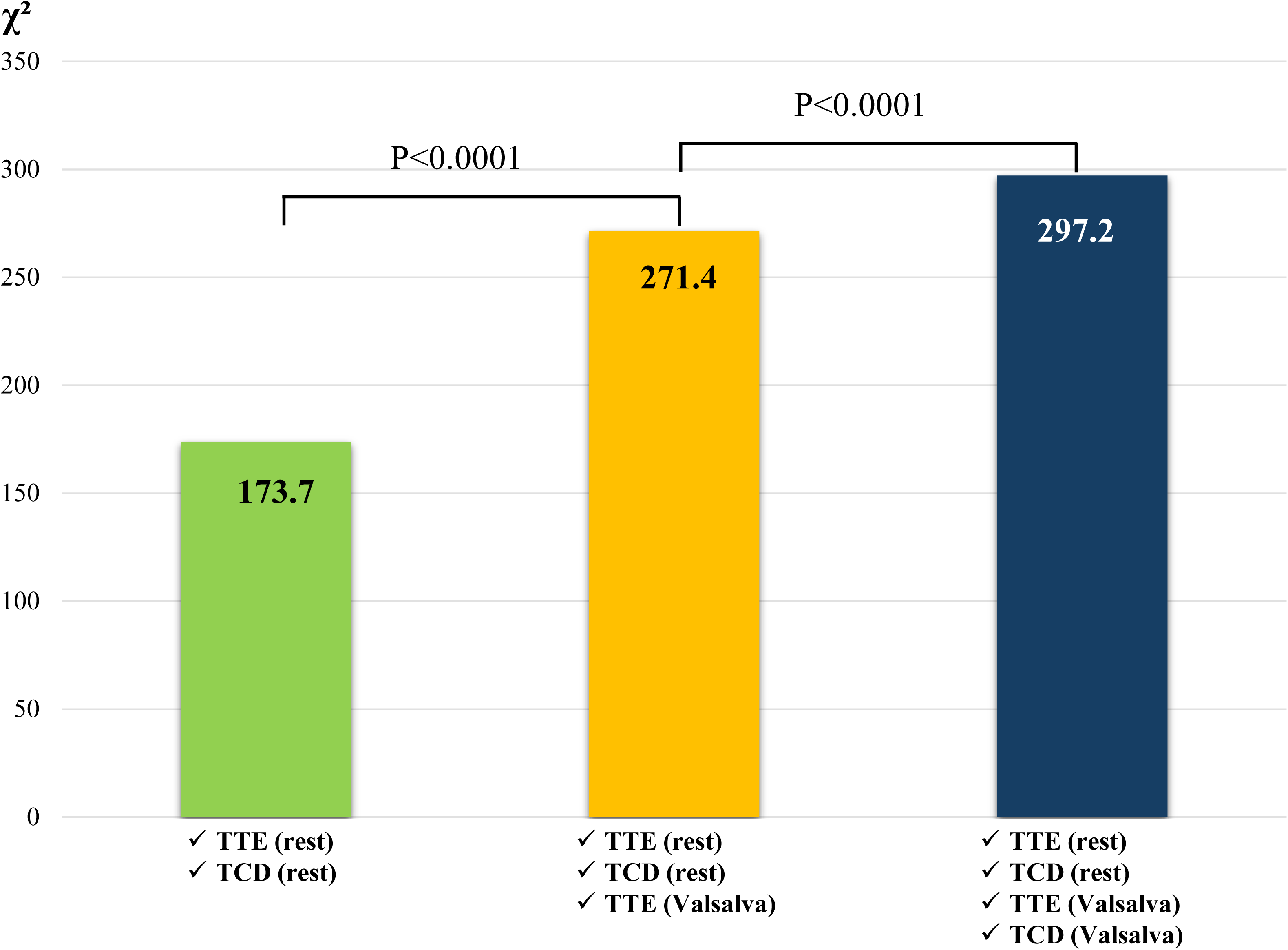
Sequential logistic regression model evaluating the incremental diagnostic value of combined screening modalities for PFO detection. A stepwise increase in model chi-square statistics was observed with the sequential addition of bubble test parameters. The baseline model incorporating TTE at rest and TCD at rest yielded a chi-square of 173.7. Inclusion of TTE under VM significantly enhanced model fit (χ²=271.4, p<0.0001), and subsequent addition of TCD during VM further improved performance (χ²=297.2, p<0.0001). TTE = transthoracic echocardiography, TCD = transcranial Doppler, VM = Valsalva maneuver

In this model, the chi-square statistic value for the combination of TTE (rest) and TCD (rest) was 173.7. The addition of TTE (VM) significantly improved model performance, increasing the chi-square value to 271.4 (p<0.0001). Incorporating TCD (VM) into this model resulted in a further significant step-up of the chi-square statistic to 297.2 (p<0.0001). These results demonstrate a significant incremental diagnostic value of TCD (VM) when added to TTE (Valsalva), indicating enhanced predictive ability for detecting PFO.

### Concordance and complementary value of TTE and TCD

To elucidate the reason why adding TCD to TTE under VM significantly enhanced predictive accuracy, even though the AUCs of TTE and TCD under VM were comparable, we analyzed the concordance of shunt grades between TTE (VM) and TCD (VM) (Table 4). The column labeled “OR” rule indicates cases that were positive (shunt grade ≥ small) on either TTE (VM) or TCD (VM) but negative on the other test. Agreement between TTE (VM) and TCD (VM) findings was assessed using Cohen’s kappa statistics. The overall concordance rate of shunt grade between TTE (VM and TCD (VM)) was 75.2% (300/399), with a kappa coefficient of 0.63 indicating substantial agreement and a quadratic-weighted kappa of 0.86 indicating strong agreement. A total of grade discordance between TTE and TCD under VM was observed in 99 of 399 cases (24.8%), among which 54 cases were diagnosed with PFO. Notably, 54 patients were positive in either TTE (VM) or TCD (VM) but negative in the other modality, thus classified as “OR”rule positive. This subgroup comprised 24 TTE (VM)-only positive and 30 TCD (VM)-only positive cases, with TEE-confirmed PFO in 15 patients (28%), including 6 in the TTE (VM)-only group and 9 in the TCD (VM)-only group. Next, Table 5 presents the distribution of true-positive, true-negative, false-positive, and false-negative results, along with the corresponding sensitivity and specificity, when a shunt grade of ≥ small was defined as positive. Compared with TTE (VM) or TCD (VM) alone, the “OR” rule (either positive) showed significantly higher sensitivity (TTE [VM] alone vs “OR”, p=0.0039; TCD [VM] alone vs “OR”, p=0.031), whereas the “AND” rule (both positive) demonstrated significantly higher specificity (TTE [VM] alone vs “AND”, p<0.0001; TCD [VM] alone vs “AND”, p<0.0001). No significant differences in sensitivity and specificity were observed between TTE (VM) and TCD (VM) (p=0.61 and p=0.75, respectively). These findings suggest that performing both TTE and TCD yields complementary gains in sensitivity and specificity, leading to improved diagnostic accuracy compared with either modality alone.

**Table 4.**
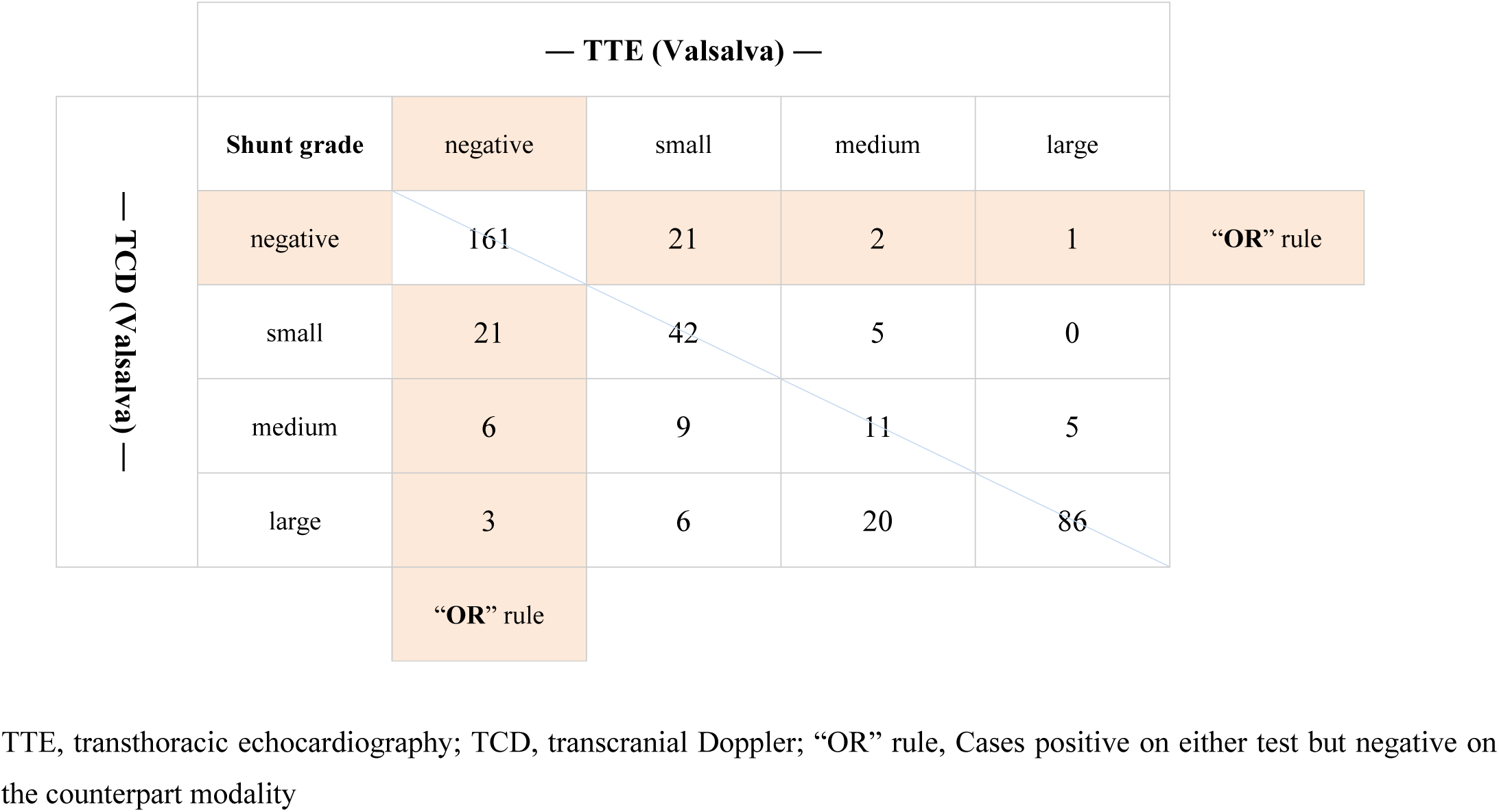
Cross-classification of TTE (Valsalva) and TCD (Valsalva) shunt grades.

**Table 5.** Comparison of dual-screening (OR/AND rules) and single tests (TTE or TCD) for detection of PFO.

| Test | TP | TN | FP | FN | Sensitivity | Specificity | p (vs TTE or TCD alone) |
| --- | --- | --- | --- | --- | --- | --- | --- |
| **TTE (Valsalva)** | 141 | 176 | 67 | 15 | 90.4% | 72.4% | N/A |
| **TCD ( Valsalva )** | 144 | 173 | 70 | 12 | 92.3% | 71.2% | N/A |
| **“OR” ( Either positive )** | 150 | 155 | 88 | 6 | 96.2% | 63.8% | Sensitivity ↑: vs TTE (p=0.0039), vs TCD (p=0.031) |
| **“AND” ( Both positive )** | 135 | 194 | 49 | 21 | 86.5% | 79.8% | Specificity ↑: vs both TTE and TCD (p<0.0001) |

TTE, transthoracic echocardiography; TCD, transcranial Doppler; TP, true positive; TN, true negative; FP, false positive; FN, false negative; N/A, not applicable; p values were calculated using McNemar’s exact test (two-tailed).

## Discussion

This study is the first to investigate, in patients with CS, the clinical significance of performing both TTE and TCD as concurrent screening modalities within a collaborative HBT framework. Incorporating TCD findings into TTE-based screening yielded a significant incremental gain in diagnostic accuracy for PFO detection. In addition, we systematically analyzed concordance and discordance patterns between TTE and TCD bubble-test grading under VM. The substantial agreement observed underscores the reliability and technical maturity of screening examinations conducted under an established HBT. Importantly, a considerable proportion of PFO-positive cases were identified among discordant results, and application of an “OR” rule (positive in either test) significantly enhanced sensitivity and reduced missed diagnoses. Although several guidelines or expert statements provide recommendations regarding either TTE or TCD individually for PFO evaluation^21, 25^, none advocate their combined or sequential use for dual screening. The present findings therefore provide novel and robust evidence supporting the complementary nature of these modalities when used in combination. Moreover, our results emphasize that the HBT framework should not be limited to decision-making for PFO closure but should also be actively involved from the diagnostic phase. Early multidisciplinary collaboration ensures both diagnostic accuracy and procedural quality, reinforcing the concept of an integrated heart–brain approach in the management of CS.

### Reappraisal of the Importance of the Valsalva Maneuver in PFO Screening

Although this study primarily focused on concurrent TTE and TCD screening, bubble-test results were systematically obtained under both resting and VM for each modality. Consistent with previous reports, the sensitivity of both TTE and TCD for PFO detection improved during VM compared with rest^14, 23, 26, 27^. ROC analyses further demonstrated that the area under the curve (AUC) was significantly greater during VM for both modalities, whereas no significant difference was observed between TTE and TCD under either condition. These findings reaffirm the critical importance of performing an adequately executed VM when conducting PFO screening using either TTE or TCD.

### Optimal Microbubble Thresholds for Minimizing Missed PFO Diagnoses

Another important finding of this study is the differential predictive performance of each shunt-grade threshold on TTE and TCD. Across both rest and VM conditions, defining positivity at the small grade consistently yielded the highest sensitivity, whereas using larger grades as the cutoff more improved specificity at the expense of sensitivity. From a screening standpoint—where minimizing missed diagnoses is paramount—our data indicate that adopting grade ≥ small as the positive threshold is most appropriate for patients with suspected CS. In contrast, a prior study reporting that “≥5 microbubbles most accurately predicts PFO” employed a considerably younger cohort (mean age ≈42 years), including many patients with migraine, in whom PFO is more likely to represent the primary causal mechanism and shunt magnitude tends to be greater^28^. In such populations enriched with large functional shunts, even a stringent threshold can maintain reasonable sensitivity, while the higher cutoff inevitably improves specificity. By comparison, the present cohort represents a more heterogeneous and clinically realistic CS population (median age 68 years), including patients with vascular risk factors such as hypertension. In such individuals, PFO—when present—may manifest with only minimal microbubble appearance. Consequently, raising the positivity threshold reduces sensitivity because small-grade shunts still contain a non-negligible proportion of true PFOs. These population differences likely account for the divergent optimal thresholds between studies and further support the use of grade ≥ small for screening in routine CS practice.

### Comparison of the Diagnostic Performance Between TTE and TCD

Teague and Sharma simultaneously performed two-dimensional contrast echocardiography and TCD in 46 patients with stroke or transient neurological symptoms, demonstrating that RLS was detected by TTE in 26% at rest and 15% during VM, whereas TCD identified RLS in 41% under both conditions, with an overall concordance of 75–82%^29^. Most discordant cases were TCD-positive, suggesting higher sensitivity of TCD for detecting paradoxical embolization. Subsequent comparative studies have also shown excellent diagnostic performance of TCD, with pooled sensitivity and specificity of 97% and 93%, respectively, in meta-analyses^30^. In contrast, the reported sensitivity of TTE has been highly variable across studies, with a pooled sensitivity of 46% and a remarkably high specificity of 99%^17^. Consistent with these findings, our present data demonstrated slightly lower sensitivity for TTE (VM) than for TCD (VM), whereas specificity remained comparable between the two modalities. These results align well with prior meta-analytic evidence, confirming that our dual-modality screening was performed under appropriately standardized conditions and yielded results consistent with established benchmarks. In our study, the high concordance of bubble-test grading between TTE and TCD during VM indicates that both examinations were performed with consistently high technical quality. However, compared with previous reports, the specificity of both modalities appeared relatively low. This discrepancy may be partly explained by the inclusion of patients with concomitant pulmonary or malignant diseases, as well as elderly individuals with multiple comorbidities, among whom the diagnostic work-up often encompassed a broad search for embolic sources. In such populations, the presence of intrapulmonary arteriovenous anastomoses (IPAVAs)—vascular channels connecting pulmonary arteries and veins that can open under certain physiological or pathological conditions—has been described as a potential source of microbubble passage independent of PFO^31–33^. Small-scale IPAVAs may not always be captured by TTE or TCD depending on vessel caliber and flow velocity, yet can occasionally be visualized as fine, delayed-appearing microbubbles on TEE. This mechanism may have contributed to the slightly increased rate of false-negative results for TTE and TCD observed in our cohort.

### Combined “OR” rule of TTE and TCD Reduces False Negatives and Enhances Sensitivity for PFO Detection

Minimizing missed diagnoses—that is, reducing false negatives to the lowest possible level—is the fundamental principle of any screening test. The major clinical implication of the present study lies in demonstrating that performing bubble tests with both TTE and TCD during VM and interpreting them according to an “OR” rule (positive if either test is positive), significantly improves the detection sensitivity for PFO. Although only a few studies have examined combined TTE–TCD screening, Yang et al. reported that the integration of both modalities improved specificity and reduced misclassification compared with either test alone. In their analysis, when only large shunts detected within three cardiac cycles on both TTE and TCD were combined using an “AND” rule, sensitivity decreased to 39%, but specificity reached 100% with no misclassification^13^. Because TEE remains indispensable for definitive diagnosis and morphological assessment, the primary role of TTE and TCD should be to maximize sensitivity and minimize false negatives. In our cohort, the “AND” rule similarly increased specificity compared with either test alone, whereas the “OR” rule significantly reduced false negatives and improved overall sensitivity at the expense of slightly more false positives. Despite each modality already possessing reasonable diagnostic performance individually, their combination acted in a complementary manner, thereby reducing missed diagnoses and validating the clinical utility of dual-modality screening. Notably, in this study, patients classified as “either positive” accounted for 14% of the entire cohort (n = 54), and 15 of these (28%) were confirmed to have PFO with RLS on TEE—cases that would have been overlooked if only one screening modality had been performed. These findings underscore that the complementary nature of dual-modality screening is not merely statistical but arises from fundamental differences between the two techniques. TTE provides direct anatomical visualization of the interatrial septum and the moment of shunt passage; however, its performance is highly dependent on thoracic morphology, respiratory mechanics, and interindividual variability in the acoustic window. Consequently, image quality may be suboptimal in elderly or obese patients, leading to difficulty in capturing microbubble-mediated RLS in a subset of cases. In contrast, TCD depends on the acoustic transmissibility of the temporal bone, and inadequate transtemporal windows are known to occur in a proportion of adults^34^. Moreover, TCD does not visualize anatomy but instead detects microbubble transit as MES. While this enables detection of subtle shunting at the level of the interatrial septum, it also renders the modality susceptible to confounding signals originating from intrapulmonary shunts or minimal background noise. These intrinsic differences in sensitivity profiles and modality-specific limitations likely account for the discordant cases observed between TTE and TCD. In turn, they provide the mechanistic explanation for why performing both tests creates a mutually compensatory framework, thereby enhancing overall diagnostic performance.

### Heart–Brain Team Collaboration at the Diagnostic Stage

The present findings underscore the practical value of implementing dual TTE–TCD screening within a structured HBT workflow. Traditionally, the HBT has been emphasized as a multidisciplinary forum for therapeutic decision-making, particularly regarding the indication for PFO closure^18–20^. However, no previous study has focused on the collaborative performance of diagnostic screening itself. Our approach demonstrates that involving both cardiologists and neurologists from the diagnostic stage enables higher-quality, pathophysiologically grounded, and unbiased assessment of potential embolic sources, thereby facilitating appropriate subsequent management. This diagnostic-phase collaboration represents a natural extension of the HBT concept and may serve as a model for comprehensive, balanced decision-making in the management of patients with CS.

### Limitations

This study has several limitations. First, it was conducted at a single center with a specific HBT protocol, which may limit generalizability. Second, the retrospective design may have introduced selection bias despite the consecutive enrollment. Third, TEE served as the reference standard, but variability in VM performance or acoustic window quality might have influenced bubble visualization. Fourth, the study population included relatively elderly patients, some of whom had intrapulmonary shunts or comorbid cardiopulmonary and malignant diseases, which could have affected the specificity of TTE and TCD. Finally, the study did not assess the prognostic impact of dual screening on clinical outcomes, which warrants future prospective validation.

## Conclusion

In patients with CS, performing both TTE and TCD bubble tests within a HBT framework significantly enhances the diagnostic sensitivity for detecting PFO and reduces missed diagnoses. Applying an “OR” rule (positive if either test is positive) during VM provides a practical and reliable strategy for initial screening. These findings highlight the value of early multidisciplinary collaboration between cardiology and neurology in achieving accurate and pathophysiologically grounded evaluation of paradoxical embolic sources.

## Acknowledgements

The authors would like to express their gratitude to the sonographers at the Ultrasound Centre of Showa Medical University Hospital for performing the transthoracic echocardiographic examinations. We also thank Editage (www.editage.jp) for professional English language editing.

## Data availability

The datasets generated and analyzed during the current study are available from the corresponding author upon reasonable request.

## Author contributions

Ken Oshima: Conceptualization, Data curation, Formal analysis, Writing—original draft. Yasuhide Mochizuki: Supervision, Conceptualization, Methodology, Writing—review and editing.

Ayaka Oda, Mamiko Yamada, Yumi Yamamoto, Sakiko Gohbara, Saaya Ichikawa-Ogura, Rumi Hachiya, Eiji Toyosaki and Hiroto Fukuoka: Echocardiographic data acquisition and image interpretation.

Keita Mizuma, Tetsuhito Nohara, Ayako Miki and Hidetomo Murakami: Clinical data collection and patient management as a stroke specialist.

Yasuhide Mochizuki: Statistical analysis, Methodology, Critical review of the manuscript.

Naoki Uchida and Toshiro Shinke: Supervision, Conceptual advice, Final approval of the manuscript.

All authors approved the submitted version of the manuscript and agreed to be accountable for all aspects of this work.

## Ethical approval

This study was approved by the local ethics committee of Showa Medical University Hospital (approval number: 2025-0407), and by University Hospital Medical Information Network (UMIN 000060079).

## Consent to participate

Informed consent of this research was obtained in the form of an opt-out on the website, and individuals who refused were excluded from the study.

## Non-standard Abbreviations and Acronyms

CS: cryptogenic stroke
PFO: patent foramen ovale
RLS: right-to-left shunt
TTE: transthoracic echocardiography
TCD: transcranial Doppler
TEE: transesophageal echocardiography
VM: Valsalva maneuver
MES: Micro-embolic signals
HBT: Heart–Brain Team
RoPE: Risk of Paradoxical Embolism
DVT: deep vein thrombosis
PE: pulmonary embolism
IPAVA: intrapulmonary arteriovenous anastomosis

